# Characteristics of mental health stability during COVID-19: An online survey with people residing in the Liverpool City Region

**DOI:** 10.1101/2022.03.17.22272479

**Authors:** Katalin Ujhelyi Gomez, Rhiannon Corcoran, Adele Ring, Shaima Hassan, Katherine Abba, Jennifer Downing, Mark Goodall, Mark Gabbay, Pam Clarke, Paul Moran, Dorcas Akeju Obe, Kate M Bennett

## Abstract

**Background and aim:** Despite the significant mental health challenges the COVID-19 pandemic and its associated government measures have presented, research have shown that the majority of people have adapted and coped well. The aim of this study was i) to determine the proportion of people with mental stability and volatility during the pandemic in a North West urban environment sample and ii) to establish group differences in psychosocial variables. Mental stability and volatility refer to the extent to which individuals reported change in levels of common mental health symptoms over the course of 12 weeks.

**Method:** a two-wave-online survey (N = 163) was used to explore the psychological and social impact of the pandemic on relatively disadvantaged neighbourhoods within the Liverpool City Region over 12 weeks. Kruskal-Wallis with post-hoc tests were used to determine how people with mental stability and volatility differed on factors categorised within an ecological framework of resilience (individual, community, societal, and COVID-19 specific).

**Results:** Individuals categorised as ‘stable’ in terms of mental health symptoms (63.6%) had better mental and physical health; were more tolerant of uncertainty; reported higher levels of resilience and wellbeing compared to ‘very volatile’ people (19.8%). These individuals also reported feeling less socially isolated, experienced a greater sense of belonging to their community which was more likely to fulfil their needs, and were more likely to have access to green space nearby for their recommended daily exercise. ‘Stable’ individuals did not report worrying any more during the pandemic than usual and tolerated uncertainty better compared to those in the ‘volatile’ group.

**Implications:** The majority of participants in this sample were mentally stable and coping well with the challenges presented by the pandemic. The resilience of these individuals was related to key place-based factors such as a strong sense of community and useable local assets. The data showcase the role of place-based social determinants in supporting resilience and thereby highlight key preventative measures for public mental health during times of international crisis.

## Introduction

The novel coronavirus SARS-CoV-2, rapidly spread across the world becoming a global pandemic by March 2020. As a response to tackle the virus and limit its spread, the UK Government initiated a national lockdown on the 23^rd^ of March 2020 that lasted for over four months. This was followed by various levels of restrictions and public health advice throughout the country for the coming months from social distancing and working from home, mask wearing and hand hygiene through to stringent domestic and international travel restrictions. The ongoing pandemic and the associated government measures have dramatically changed the way people live, work, and socialise ^1^.

Several research groups have reported that the coronavirus pandemic has presented significant mental health challenges ^2-5^. In line with findings in other countries (e.g. ^6,7^), the Office of National Statistics (ONS) have reported elevated distress indices, such as increased levels of anxiety (32%), diminished well-being (43%), and loneliness (23%) among the UK population ^8^. However, some studies have identified different trajectories for mental health (e.g. ^9,10^). For example, Shevlin et al. (2021) found five classes reflecting stability (low stable and high stable), improvement and two classes of deterioration (one more severe than the other). The most common trajectory was a low-stable profile, which could be termed resilient. A systematic review and meta-analysis of 65 pandemic mental health cohort studies suggest that the initial increase in mental health symptoms resolved towards the norm with time, which has been suggested to demonstrate the impact of resilience ^11^.

### Resilience

Resilience has been defined as a psychological trait which promotes well-being ^12^ or a process ^13^ leading to an outcome ^14^. Windle (2011) ^13^, following from a comprehensive concept analysis, defines resilience as “*the process of effectively negotiating, adapting to, or managing significant sources of stress or trauma. Assets and resources within the individual, their life and environment facilitate this capacity for adaptation and ‘bouncing back’ in the face of adversity*.*”* ^13^. Resilience can also be considered as a capacity for mental stability or toughness ^15^ through adapting and bouncing back in the face of challenge ^14^, protecting against stress, trauma, and adversity ^16^. However, there are wider determinants of resilient responses to negative situations and trauma ^17^. To account for the interaction between different levels of factors that can influence a person’s resilience, an ecological model of resilience has been proposed ^18^. This framework (Fig 1) recognises that resilience operates interactively in context at individual, community, and societal levels ^13,19^. Individual resilience thus develops amidst an interplay of environmental-community and social-political factors, most of which are beyond individual control ^20^.

### Resilience and COVID-19

Higher resilience has been linked to reduced psychological symptomatology ^21^ and distress ^22^, as well as increased wellbeing ^23-25^. Preliminary data indicates that the pandemic has increased people’s acute and chronic stress levels due to the uncertainty and uncontrollability of the pandemic’s impact on broad aspects of autonomy, health, family and finances ^26^. COVID-related resilience has been negatively associated with symptoms of depression, anxiety, somatisation, and negative emotional symptoms ^27^. People have been shown to cope with the pandemic through connections with loved ones, going outdoors, physical activity, via spirituality ^28,29^, leading a healthy lifestyle, accepting anxiety and negative emotions, and inquiring information about medical treatment ^30^. In a test of the ecological model of resilience during the pandemic in Italy, evidence supporting the role of individual, societal and COVID-specific levels of the model (but not community), was found. Key contributors to resilience in this study included: psychological variables such as conscientiousness and intolerance of uncertainty; demographic variables such as having children in the home and educational level; and covid-specific variables such as COVID-specific anxiety, and social distancing ^31^.

In the UK, the COVID-19 crisis has had a particularly negative effect with one of the highest mortality rates in the world ^32^. Uncertainty is always challenging and can contribute to the development of generalised anxiety, depression, and health anxiety ^33^. A study of 555 UK adults ^34^ observed high levels of generalised anxiety (27%) as a result of the pandemic that was more than four times the national average pre-pandemic 5.9%; ^35^. The figure was also higher compared to rates identified by previous pandemic research (e.g. ^36,37^).

However, increased anxiety, loneliness, distress and low mood are normative experiences during such a crisis and it is one’s response to these negative emotions that ultimately influence longer-term mental health outcomes ^38^. Individual responses to stress are affected by coping style, available social support, previous experience with the specific stressor, underlying mental health issues, and personality characteristics ^26^. While the negative mental health impact of the pandemic is real, generally, adaptation and recovery are the typical responses to trauma and adversity ^39^. This is apparent in research findings among the UK population collected during the pandemic. In spite of struggles as the crisis unfolded in the UK, 64% of people who responded to a national longitudinal survey reported coping well ^10^, using adaptive coping strategies, such as going for a walk, spending time in open spaces, staying connected with others, or exercising ^40,10,41^. Another study found the prevalence of psychological problems at the early stage of the crisis only slightly higher than during previous pandemics suggesting again the population’s successful adaptation to the situation ^42^. As mentioned previously, in a longitudinal study of more than 2000 participants, Shevlin et al. (2021) ^9^ identified five profiles (low stable; high stable; improving; and two deteriorating profiles). They found that psychological variables distinguished between the low-stable and the other profiles.

Research that helps us understand the wide range of determinants that impact on individual ability to respond resiliently can facilitate evidence-based policy management of future or ongoing crises. Moreover, while research has been conducted on a national level, in-depth localised research in areas where the pandemic hit harder is relatively lacking.

### The aim of the current study

In order to fill the above research gaps, the current study adopted a resilient systems approach to understand individual responses to the pandemic. Additionally, it investigated the psychological and social impact of the COVID-19 pandemic and its associated lockdown restrictions in a city region located in the North of England with high levels of deprivation and thus higher level of vulnerability to the pandemic.

Using Windle and Bennett’s (2011) ^18^ ecological model of resilience, this study focused on demographic, psychological, community, and societal factors that may determine individual response to the pandemic. Additionally, COVID-19 specific factors were included to assess their capacity to influence resilience. In this study, an individual’s resilience during the pandemic was defined as the extent to which they reported change in levels of common mental health symptoms over the course of 12 weeks. Mental stability may reflect a person’s adaptation to and coping with challenges, while more vulnerable people may be more prone

to volatility or change in mental health. The study aims to establish i) the proportion of people with mental stability and mental volatility and ii) differences between groups in terms of demographic, psychological, societal, community, and COVID-specific factors.

## Methods

### Design

These data are part of a larger study surveying households early in the COVID-19 pandemic and then twice more in three months. The study explored the psychological and social impact of the pandemic and its associated restrictions on relatively disadvantaged neighbourhoods within a city region in the North West, and factors influencing response to and impact of the pandemic. An online survey using JISC software was launched in mid-June 2020 and conducted three times, across a 12-week period (week 1, week 6, week 12).

### Participants and recruitment

Participants were recruited using several methods including re-contacting, by telephone or mail, willing participants who had supplied data for either or both wave 1 or wave 2 of the NIHR CLARHC North West Coast Household Health Survey (CLAHRC NWC HHS) ^43^ and by advertising the research via local media (newspaper, radio) and social media (Twitter, Facebook). The first phase of the survey collected data from 290 LCR residents.

Baseline responses were collected between mid-June and the end of August 2020. While the survey included 3 waves, the data collected at week 6 was a much-abbreviated set of questions focussing on behaviours and pastimes only. The analyses reported here uses the fuller data collected in the first and third waves, representing 12 weeks of individual pandemic experience between mid-June and mid-December 2020. Automated reminders to complete the current wave of the survey were generated through the JISC software.

Ethical approval was granted by the University of Liverpool Central Research Ethics Committee (7739). Informed consent was collected at the beginning of the first survey. Non-completion of this survey and follow-up surveys was treated as withdrawal of consent such that only data from individuals who reached the end of the survey was included in this dataset. As a result, missing data was minimal.

### Data collection and measures

This extensive survey assessed a wide range of psychological and social determinants. For the full list of measures used refer to Supplemental Table (ST) 1. This study focuses on a subset of the variables (Fig 2). The first survey comprised multiple measures assessing demographic and socioeconomic factors, mental health and wellbeing, and characteristics at the individual, community, and societal levels. The week 12 follow-up included all questions in the first survey with the exception of the psychological trait measures which would not be expected to change over time (e.g. Intolerance of Uncertainty). Fig 2 includes the variables used here within the Ecological Model of Resilience.

### Mental Health Stability Outcome Variables

The 9-item <u>*Patient Health Questionnaire*</u> (PHQ-9) ^44^ and the <u>7-item *Generalised Anxiety Disorder* (GAD 7)</u> questionnaire ^45^ were used to measure self-reported common mental health symptoms. On both measures, responses range on a four-point Likert scale from 0 ‘not at all’ to 3 ‘nearly every day’. The higher the total score, the more severe the symptoms. Both scales measure symptoms over the past week in this survey. The PHQ-9 is a valid and reliable measure (Cronbach’s α=0.86) not only for the identification of depression but also to measure its severity which makes it ideal to track changes in depression levels over time ^44^. Similarly, GAD-7 is a valid and efficient tool (Cronbach’s α=0.91) to screen anxiety and its severity in clinical practice and research ^45^.

As symptoms of depression and anxiety are typically co-morbid and with a view to meaningfully simplifying subsequent analysis, the GAD-7 and the PHQ-9 scores were summed to create a single common mental health (CMH) variable for the week 1 and week 12 data. The degree of stability or volatility of this CMH variable between week 1 and week 12 provides the working measure of individual resilience through 12 weeks of the pandemic. More detail on this derived variable is presented in the data analysis section below.

#### Individual Level Mechanisms: Demographic and sociodemographic

Demographic characteristics measured included age, gender, marital status, and accommodation. Sociodemographic variables collected were level of education, subjective assessment of financial status before the pandemic; current subjective financial status; work status before the pandemic and current work status.

#### Individual Level Mechanisms: Psychological

The 7-item <u>(SWEMWBS)</u> ^46^ assesses self-reported subjective psychological wellbeing over the previous two weeks (Cronbach’s α=0.91). Responses included 1 = ‘None of the time’, 2 = ‘Rarely’, 3 = ‘Some of the time’, 4 = ‘Often’, 5 = ‘All of the time’.

The 6-*item <u>Brief Resilience Scale</u>* <u>(BRS)</u> ^47^ measures the self-reported ability to bounce back or to recover from stress (Cronbach’s α=0.80-0.91). Participants were asked to indicate the extent to which they agreed with each statement on a scale of 1 to 5 where 1 = strongly disagree, 2 = disagree, 3 = neutral, 4 = agree, 5 = strongly agree. Items 1, 3, and 5 of the scale are positively worded, and items 2, 4, and 6 are negatively worded. The BRS was used to test construct validity of the common mental health change outcome variable, anticipating that the ‘stable’, ‘volatile’ and ‘very volatile’ groups would differ significantly with respect to their scores on the BRS.

The 12-item two-factor (prospective and inhibitory anxiety) version ^48^ of the original 27-item <u>Intolerance of Uncertainty Scale</u> (IUS) ^49^ was used to quantify emotional, cognitive, and behavioural reactions to ambiguous situations (Cronbach’s α=0.91). Response options ranged from 1 = ‘not at all characteristic of me’, 2 = ‘a little characteristic of me’, 3 = ‘somewhat characteristic of me’, 4 = ‘very characteristic of me’, 5 = ‘extremely characteristic of me’.

Participants were also asked whether they had any children, about their health status on a scale between 1 and 100 (where 100 represented the very best of health), if they had felt lonely in the previous week, and about the frequency and type of health service use in the previous 12 weeks.

#### Social/ Community Mechanisms

*The <u>Brief Sense of Community Scale</u>* ^50^ is an 8-item sense of community scale representing factors of needs fulfilment, group membership, influence and emotional connection to neighbourhood. The scale incorporates a five-point self-report Likert scale with end points from strongly agree to strongly disagree.

Additionally, respondents were asked whether they had access to a pleasant local green/open space for their recommended daily exercise; if they felt isolated from others and,

whether they would feel comfortable/uncomfortable asking a neighbour to collect a few shopping essentials for them if necessary. Participants also provided answers regarding whether they surfed the Internet more or less often than usual, or about the same as before the pandemic.

### COVID-19 specific Mechanisms

Participants were asked whether they had been or were volunteering to support their local coronavirus action; whether they had experienced worry, and whether they had felt anxious about their work situation (or the work situation of others close to them) and / or about the future.

### Data analysis

For the data analyses, variables were recoded to overcome some limitations posed by low cell sizes. See ST 2 for details.

### The COVID Resilience Grouping variable

The depression and anxiety scores on the PHQ-9 and GAD-7 for week 1 (N=163) and week 12 (N=162 due to one missing score on the GAD-7) were combined to create a composite ‘common mental health’ (CMH) variable (N=162). A ‘CMH change’ variable was then created (N=162) to reflect change in reported common mental health symptoms over the 12 weeks. This was used to create a three-level variable to capture stability versus volatility (stable, volatile, and very volatile cases) using the standard deviation of the ‘CMH change’ variable (SD=8.17) where ‘stable’ people represented those whose scores fell between -4 and +4 (no change in mental health), the ‘volatile’ group included people whose scores fell between -4 and -8, and +4 and +8 (some change in mental health), and the scores of ‘very volatile’ individuals were anywhere below -8 and above +8 (greater change in mental health).

### Statistical analyses

Kruskal-Wallis tests were used to analyse whether ‘stable’, ‘volatile’, and ‘very volatile’ individuals differed in respect of CMH at week 1 and at week 12 and BRS score at week 1. Follow-up comparisons (Mann-Whitney U tests) used Bonferroni adjustment to control for Type I error associated with conducting multiple tests. Kruskal-Wallis with post hoc tests were conducted to investigate if ‘stable’, ‘volatile’ and ‘very volatile’ participants differed in terms of their demographic, psychological, community, societal, and COVID-19 specific characteristics. Finally, Mann-Whitney U test and chi-square test of independence compared those who did and did not provide follow-up information at week 12. The analyses were conducted in IBM SPSS Statistics version 26.

## Results

### Characteristics of the sample

Participant characteristics are reported in *Tables 1, 2*, and *3*. Data was available for 163 people at both waves (baseline and 12 weeks’ follow-up). The mean age was 51.50 ranging between the ages of 22 and 84. 65% of the sample was female and participants were predominantly from white British background (95.7%). Over half of the participants (63.2%) were married or co-habiting; were educated to degree level or above (70.6%); lived in a house/bungalow (82.2%), resided in Liverpool (52.8%), with third living in the most deprived quintile, and worked full-time (50.3%) before the pandemic with 38.8% furloughed or unemployed/out of work during the crisis.

**Table 1.** Participant demographic and sociodemographic characteristics.

| Demographic and sociodemographic characteristics |  | N (%) |
| --- | --- | --- |
| **Age** |  | 163 (100%) |
|  |  | Mean age (SD) = 51.50 (14.34) |
| **Gender** |  |  |
|  | *Male* | 47 (28%) |
|  | *Female* | 106 (65%) |
|  | **Total** | 158 |
| **Ethnicity** |  |  |
|  | *Non-white* | 6 (3.7%) |
|  | *White* | 156 (95.7%) |
|  | **Total** | 162 |
| **Marital status** |  |  |
|  | *Single/Separated/Divorced/Widowed* | 55 (33.7%) |
|  | *Married/registered partnership/co-habiting* | 103 (63.2%) |
|  | **Total** | 158 |
| **Education** |  |  |
|  | *No qualification/GCSE/A level* | 46 (28.3%) |
|  | *Undergraduate/postgraduate degree* | 115 (70.6%) |
|  | **Total** | 161 |
| **Accommodation** |  |  |
|  | *House or bungalow* | 134 (82.2%) |
|  | *Flat/room* | 28 (17.2%) |
|  | *(Self-contained flat, maisonette, or apartment/ Room or rooms)* |  |
|  | **Total** | 162 |
| **Residing area within the LCR** |  |  |
|  | *Halton* | 8 (4.9%) |
|  | *Knowsley* | 15 (9.2%) |
|  | *Liverpool* | 86 (52.8%) |
|  | *Sefton* | 33 (20.2%) |
|  | *St Helens* | 6 (3.7%) |
|  | *Wirral* | 15 (9.2%) |
|  | **Total** | 163 |
| **IMD\*** |  |  |
|  | *≤ 8.49* | 7 (4.3%) |
|  | *8.5 to 13.79* | 35 (21.5%) |
|  | *13.8 to 21.35* | 30 (18.4%) |
|  | *21.36 to 34.17* | 23 (14.1%) |
|  | *≥ 34.18* | 54 (33.1%) |
|  | **Total** | 149 |
| **Work status before the pandemic** |  |  |
|  | *Full-time employed/Self-employed* | 82 (50.3%) |
|  | *Part-time employed* | 30 (18.4%) |
|  | *Full-time student/ Part-time student* | 6 (3.7%) |
|  | *Unemployed/Housewife/housebound* | 35 (21.5%) |
|  | Total | 153 |
| **Work status currently** |  |  |
|  | *Working as normal*<br>(Key worker/working in the workplace) | 33 (20.3%) |
|  | *Working from home*<br>(Employed) | 67 (41.1%) |
|  | *Furloughed/unemployed*<br>(including unemployed and claiming benefits/not working?) | 63 (38.8%) |
|  | Total | 163 |

\*IMD: Index of Multiple Deprivation Quintile group (where $\geq 34.18$ is most deprived 20% of Lower-layer Super Output Area, 2019);

**Table 2.** Participant characteristics at week 1 (continuous variables).

|  | Sample with follow-up<br>data<br>Mean (SD) | Total N |
| --- | --- | --- |
| **Individual characteristics** |  |  |
| Intolerance of uncertainty | 32.21(9.56) | 163 |
| Loneliness | 2.10(1.20) | 163 |
| Subjective health | 78.69(17.15) | 163 |
| Well-being (SWEMWBS) | 21.87(4.18) | 162 |
| Depression (PHQ-9) | 5.77(5.87) | 163 |
| Anxiety (GAD-7) | 4.61(4.97) | 163 |
| Resilience (BRS) | 3.35(0.84) | 163 |
| **Community characteristics** |  |  |
| Social isolation | 2.60(1.32) | 163 |
| Sense of community (BSCS) | 26.87(6.51) | 162 |

**Table 3.**
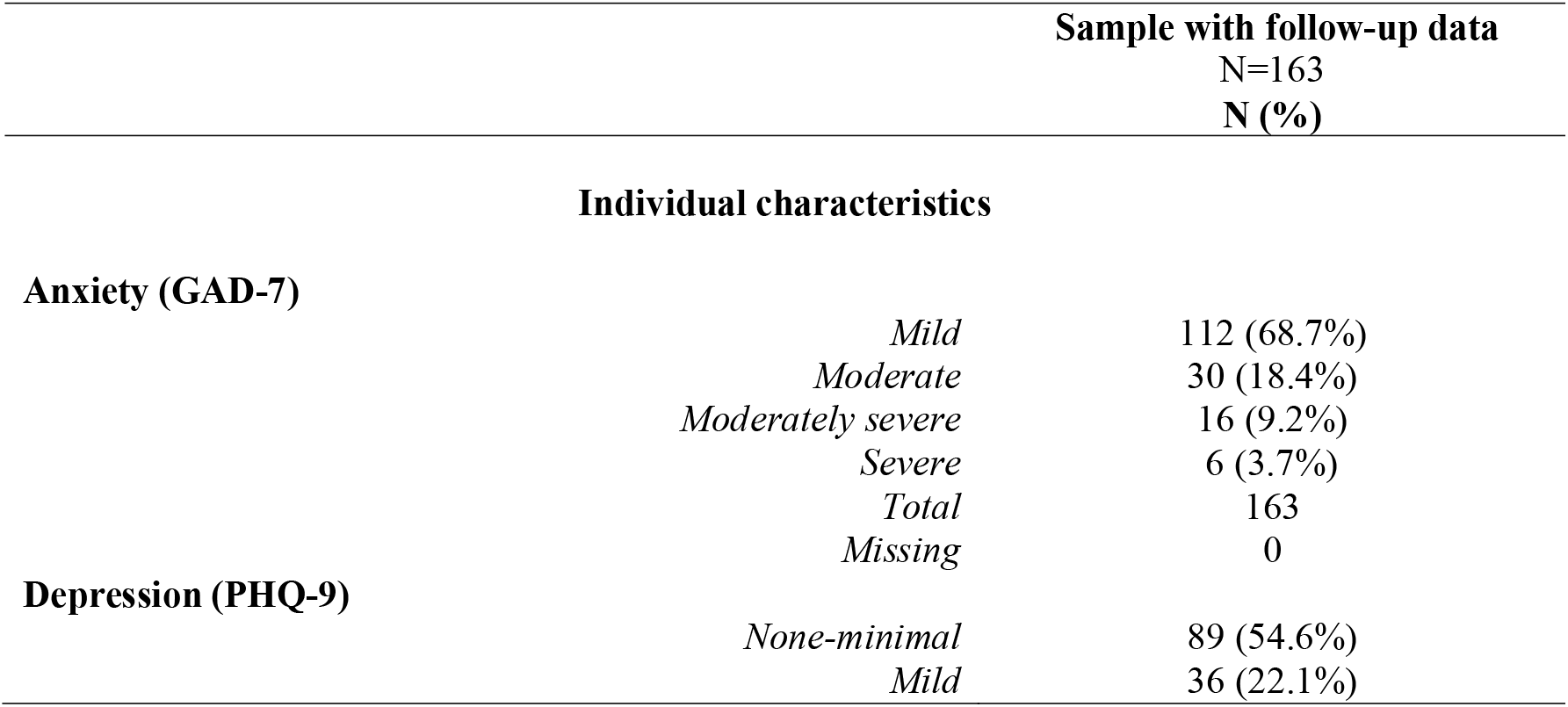
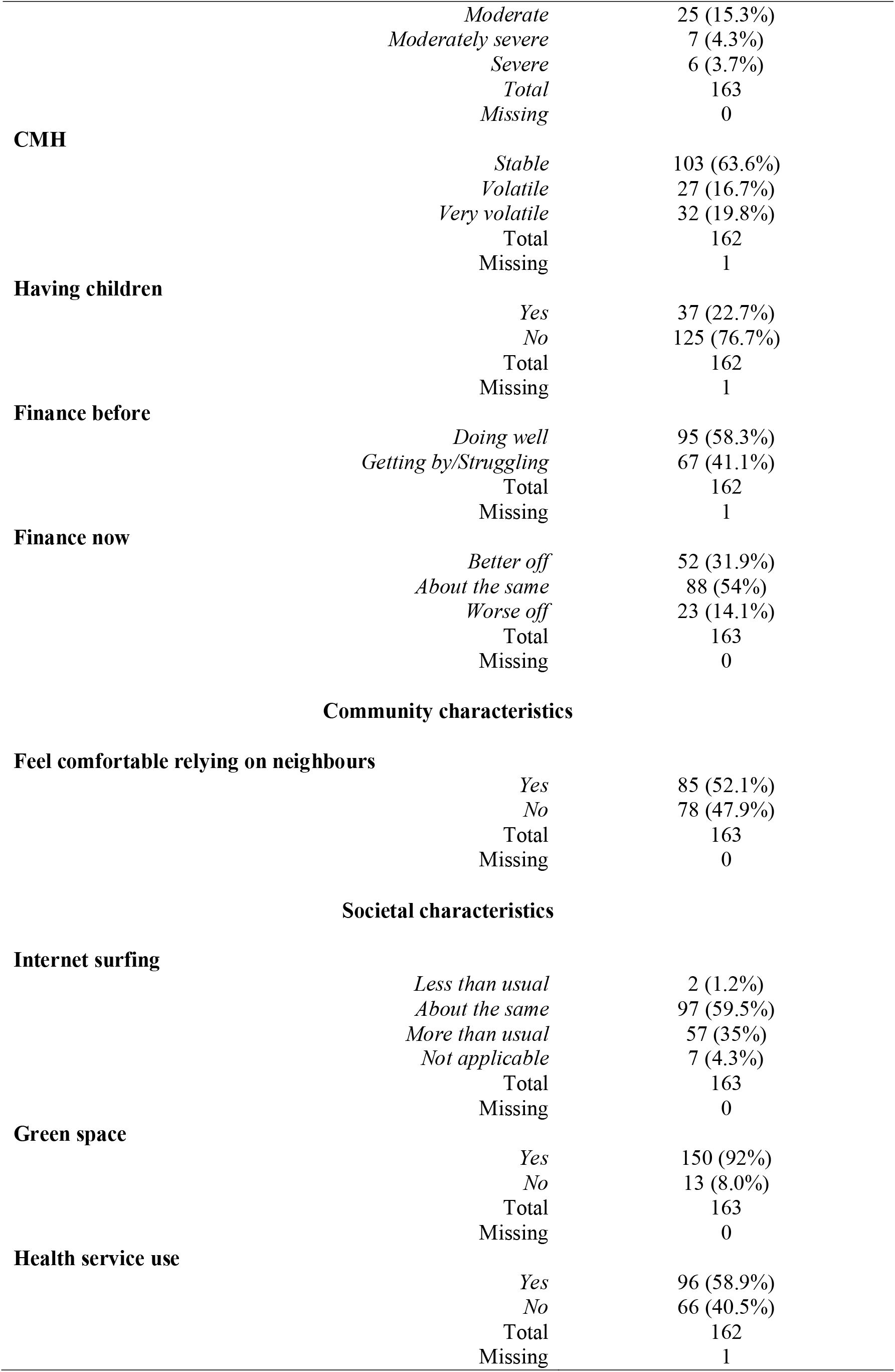
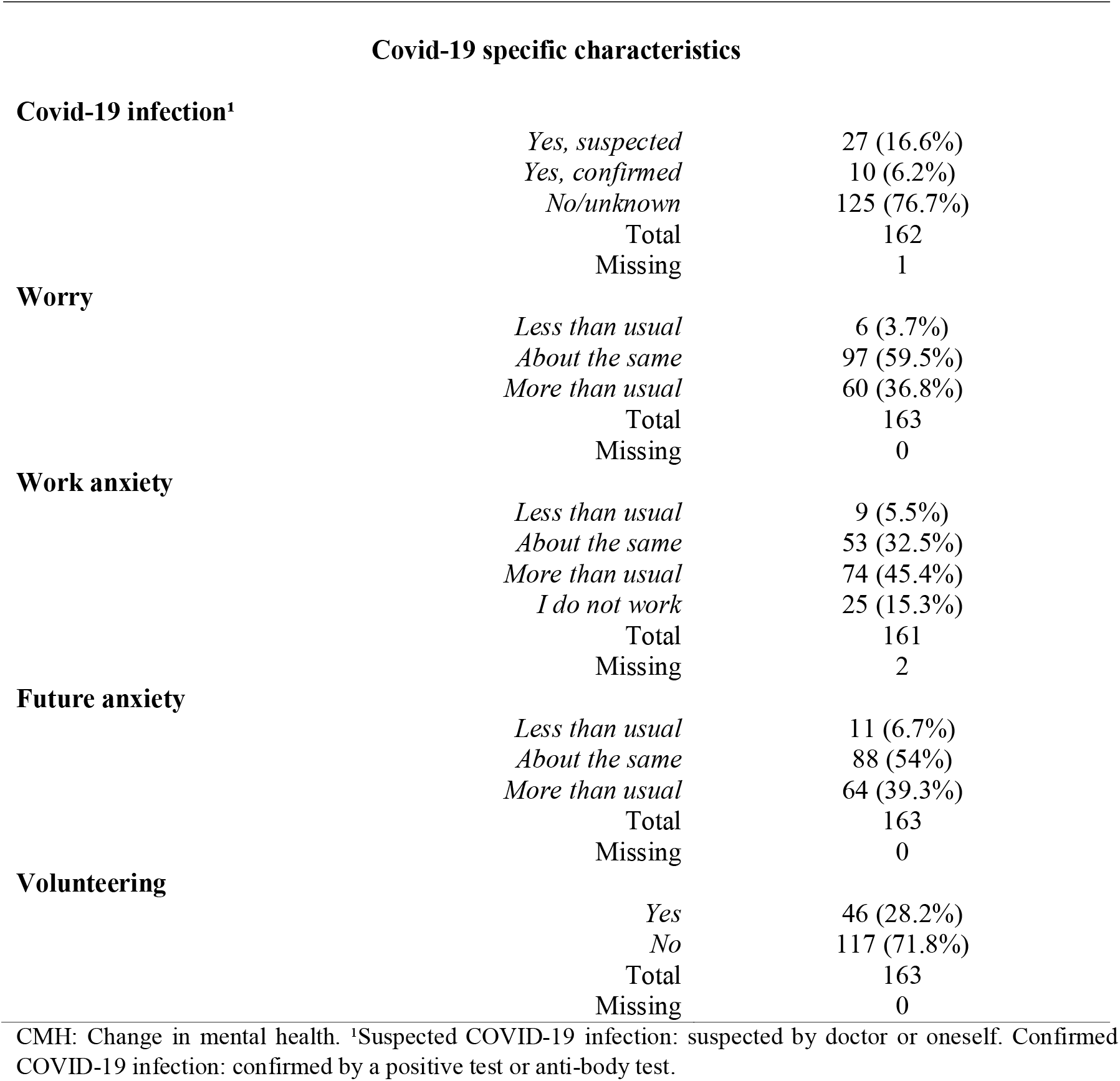
Participant characteristics at week 1 (categorical variables).

The proportion of people reporting moderate, moderately severe, or severe anxiety (≥ 10) at baseline was 35.8% in the full sample (N=290) and 31.3% in the sample with available follow-up data (N=163) at week 1. Based on changes in mental health (depression and anxiety), the 162 valid cases consisted of 103 ‘stable’ (63.6%), 27 ‘volatile’ (16.7%), and 32 ‘very volatile’ (19.8%) individuals.

### Attrition

A total of 127 people was lost to follow-up at week 12. Mann-Whitney U test showed a significant difference in age between those who completed the week 12 follow-up survey (Md=52, n=162) and those who did not (Md=46, n=125), U=8408, z=-2.54, p=.01, r=0.15, with younger people being more likely to discontinue. People without a degree were less likely to participate in the follow-up survey, χ² (1, n=287) = 13.84, p=.00, phi=.23 as demonstrated by a chi-square for independence with Yates Continuity Correction.

### Stability versus volatility

#### Mental health and resilience

The ‘Stable’, ‘volatile’, and ‘very volatile’ groups differed in respect of mental health at week 1 and at week 12. Although there was no significant difference in mental health between the ‘volatile’, and ‘very volatile’ groups, both had significantly worse mental health than their ‘stable’ counterparts at both timepoints. Mental stability/volatility was also associated with resilience levels. The mentally stable group scored significantly higher on resilience measured by the BRS compared to those in the ‘very volatile’ group. These findings remained significant at Bonferroni adjusted level (p=.017). *Table 4* provides the statistical details.

**Table 4.** Differences between the ‘stable’, ‘volatile’, and ‘very volatile’ groups in terms of mental health and resilience. Significant differences are bolded.

| | $\chi^2$<br>(p value) | Multiple<br>comparisons | N (%) | Md | Range | Z<br>score | P value<br>(effect size r) |
| --- | --- | --- | --- | --- | --- | --- | --- |
| **CMH week 1** | **37.81(0.00)** |  | 162(100) |  |  |  |  |
| Stable |  | Volatile | 27(16.7) | 14.00 | 40.00 | -0.83 | 0.41(0.11) |
|  |  | Very volatile | 32(19.8) | 18.50 | 39.00 |  |  |
| Volatile |  | **Stable** | 103(63.6) | 4.00 | 35.00 | -5.26 | **0.00\*\***<br>**(0.45)** |
|  |  | **Very volatile** | 32(19.8) | 18.50 | 39.00 |  |  |
| Very volatile |  | **Stable** | 103(63.6) | 4.00 | 35.00 | -4.26 | **0.00\*\***<br>**(0.37)** |
|  |  | **Volatile** | 27(16.7) | 14.00 | 40.00 |  |  |
| **CMH follow-up** | **33.48(0.00)** |  |  |  |  |  |  |
|  |  | Volatile | 27(16.7) | 12.00 | 37.00 | -0.36 | 0.72(0.05) |
|  |  | Very volatile | 32(19.8) | 17.00 | 44.00 |  |  |
|  |  | **Stable** | 103(63.6) | 5.00 | 39.00 | -4.40 | **0.00\*\***<br>**(0.38)** |
|  |  | **Very volatile** | 32(19.8) | 17.00 | 44.00 |  |  |
|  |  | **Stable** | 103(63.6) | 5.00 | 39.00 | -4.67 | **0.00\*\***<br>**(0.41)** |
|  |  | **Volatile** | 27(16.7) | 12.00 | 37.00 |  |  |
| **BRS Resilience** | **18.64(0.00)** |  | 162(100) |  |  |  |  |
| Stable |  | Volatile | 27(16.7) | 3.33 | 3.33 | -1.34 | 0.18(0.17) |
|  |  | Very volatile | 32(19.8) | 2.92 | 2.83 |  |  |
| Volatile |  | **Stable** | 103(63.6) | 3.50 | 3.67 | -3.79 | **0.00\*\***<br>**(0.33)** |
|  |  | **Very volatile** | 32(19.8) | 2.92 | 2.83 |  |  |
| Very volatile |  | Stable | 103(63.6) | 3.50 | 3.67 | -1.51 | 0.13(0.13) |

CMH: Common mental health. \*\*Significant after Bonferroni adjustment, $p < 0.017$ .

### Demographic factors

No significant group differences were identified in terms of demographic factors.

#### Individual factors

The analysis revealed statistically significant differences across ‘stable’, ‘volatile’ and ‘very volatile’ people in terms of intolerance of uncertainty, subjective health, and wellbeing. Further analyses among the groups revealed that ‘stable’ people reported being significantly more tolerant of uncertainty compared to ‘volatile’ and ‘very volatile’ people. This group also reported statistically significantly higher levels of subjective health, and wellbeing compared to ‘very volatile’ people.

#### Community factors

There was a significant association in relation to social isolation among the groups with the ‘stable’ group being significantly less likely to be socially isolated than ‘very volatile’ individuals. Significant differences were also found regarding sense of community, more specifically, in level of needs fulfilment and community membership. Those with stable mental health over time reported a significantly higher level of needs fulfilment from, and membership of, their community as measured by the BSCS compared to their ‘very volatile’ counterparts. These findings remained significant after Bonferroni adjustment (p<.017).

#### Societal factors

Access to a pleasant local open or green space for recommended daily exercise was associated with stability. The ‘stable’ group was more likely to have access to appropriate green space than both ‘volatile’ and ‘very volatile’ people, but the association after Bonferroni adjustment (p<.017) only remained between ‘stable’ and ‘very volatile’ people.

#### COVID-19-specific factors

There was a statistically significant difference in coronavirus-related worry across the three groups. A significantly higher proportion of the ‘stable’ group reported unchanged levels of worrying compared to ‘volatile’ and ‘very volatile’ groups with the association between stable and ‘volatile’ people remaining significant after adjustment (p<.017). Fig *3* provides an overview of the results embedded in the resilience framework and *Tables 5* and *6* include further information on these results.

**Table 5.**
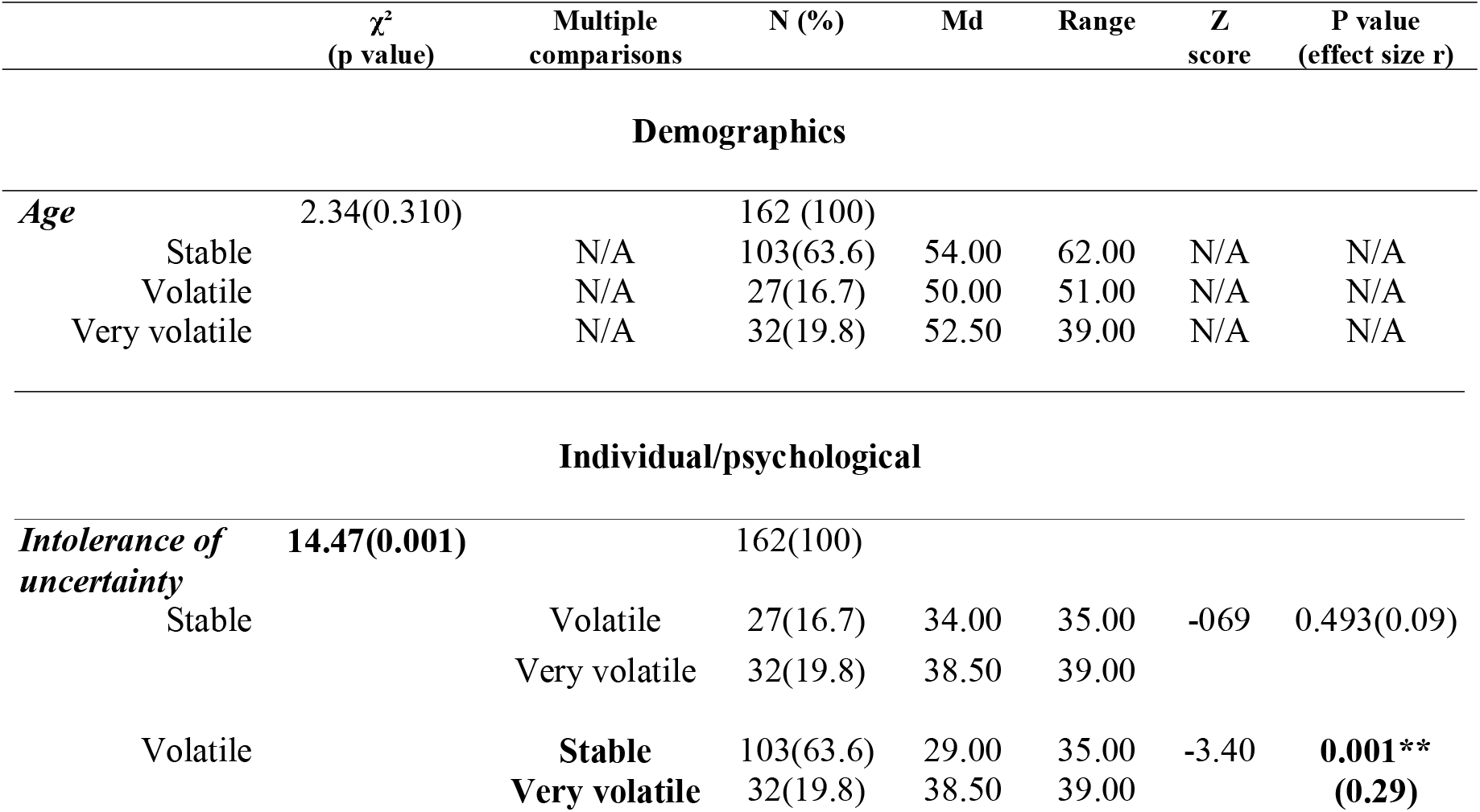
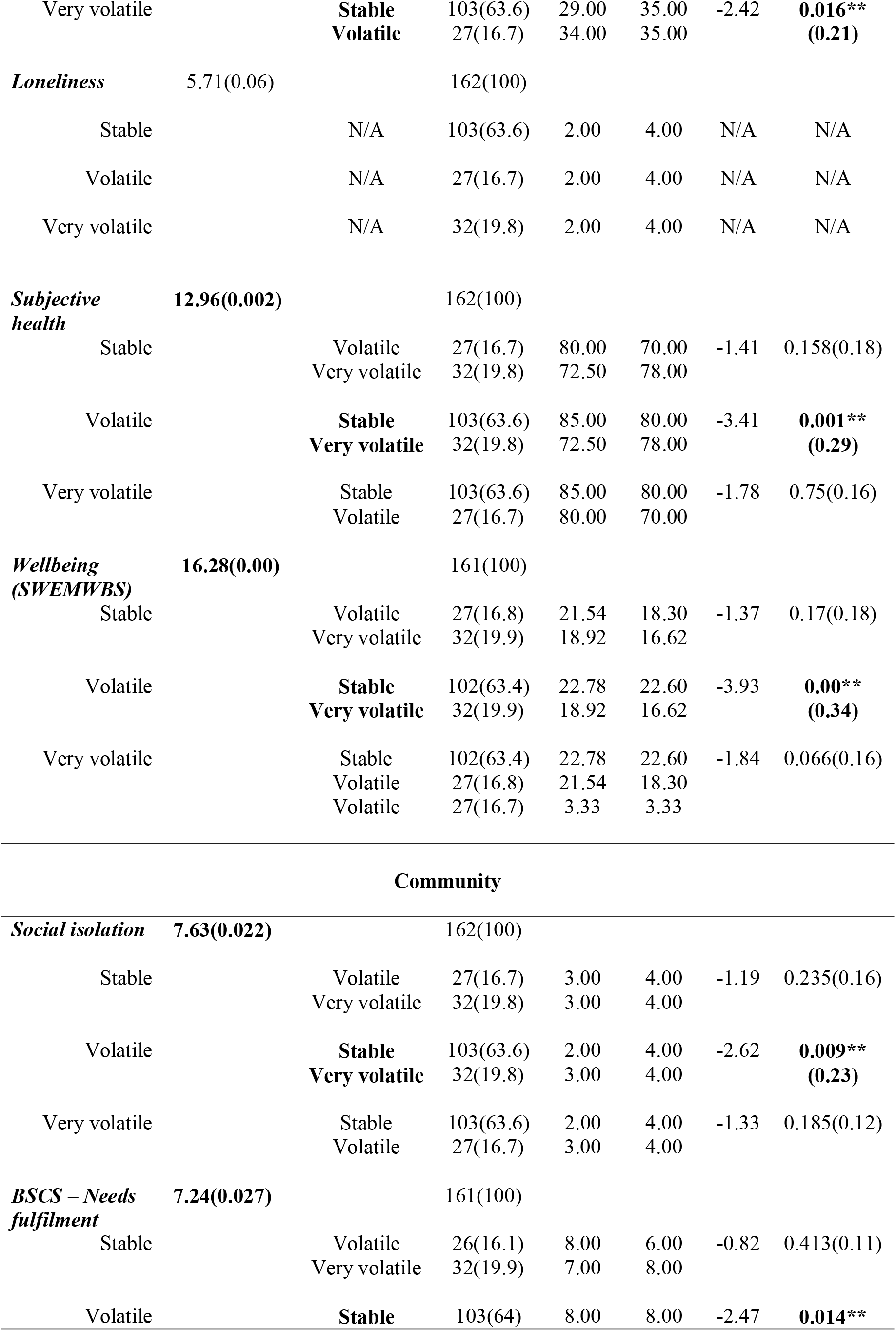
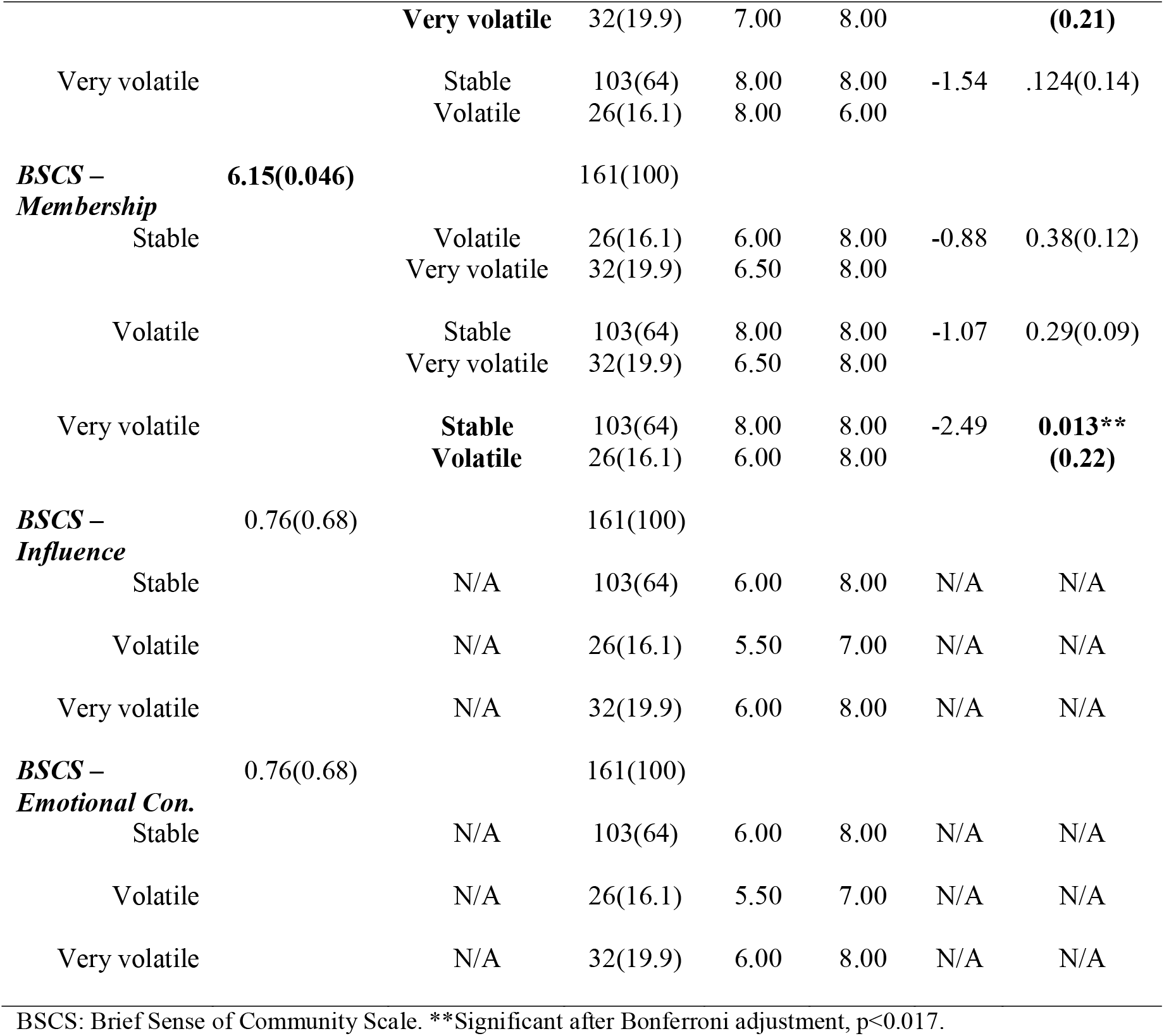
Differences between the ‘stable’, ‘volatile’, and ‘very volatile’ groups using Kruskal-Wallis and Mann-Whitney U tests. Significant differences are bolded.

**Table 6.**
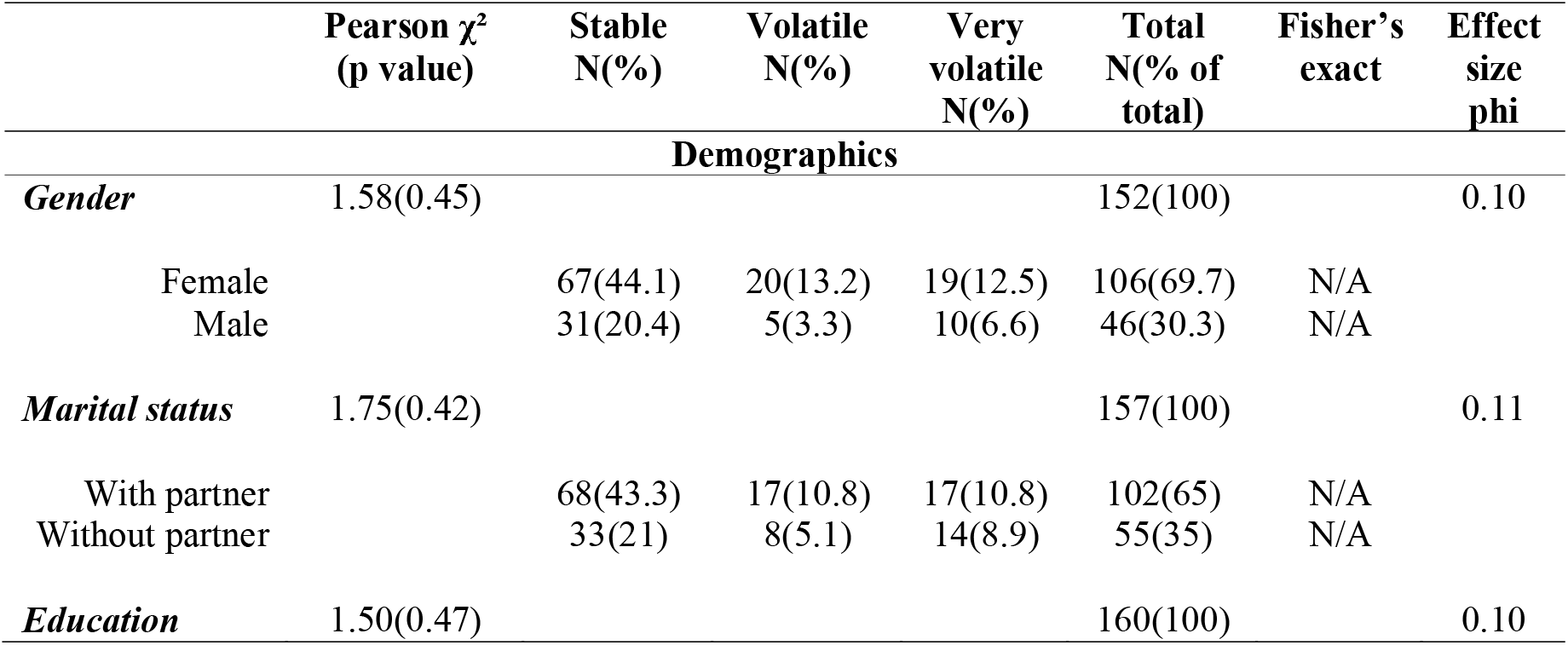
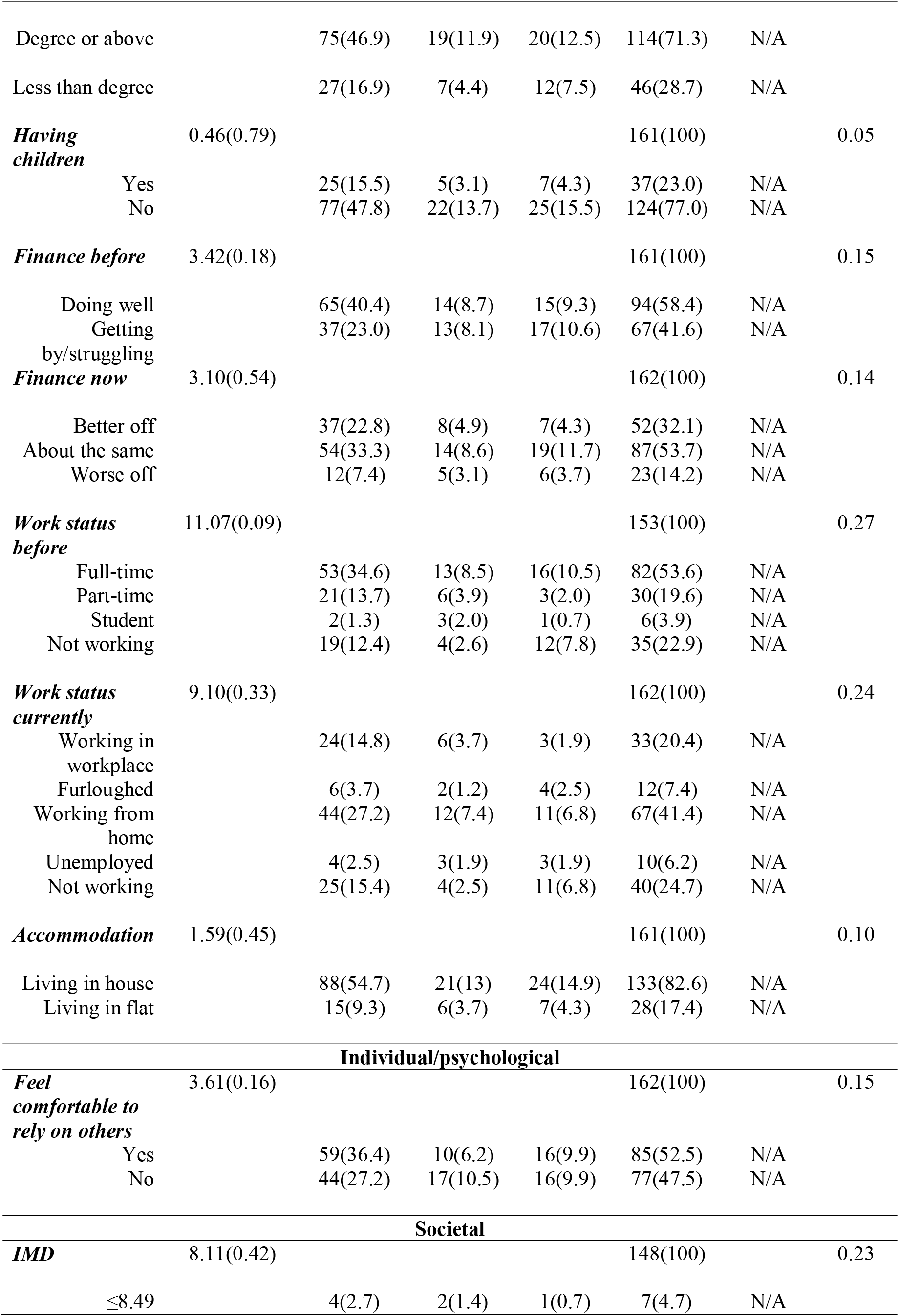
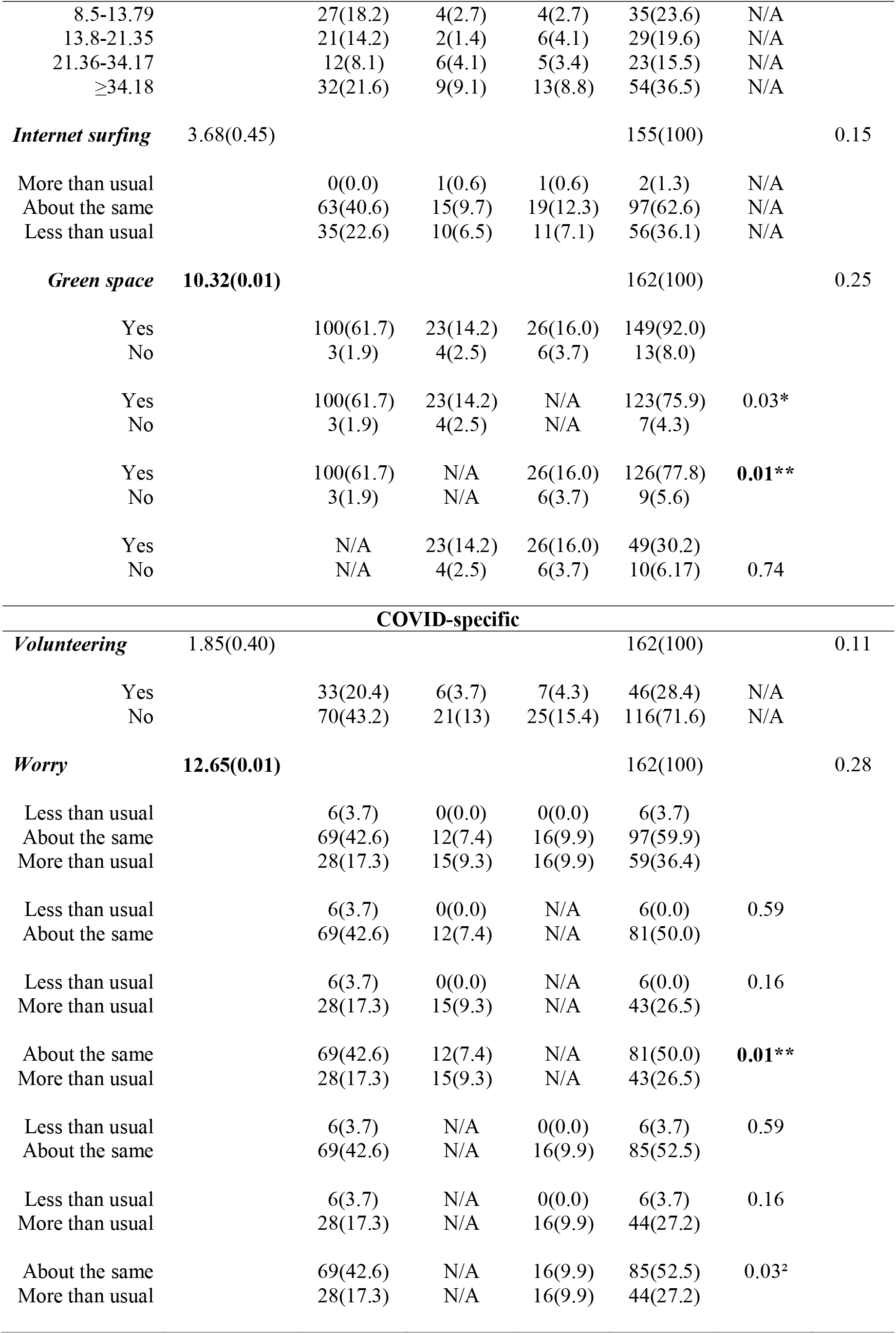
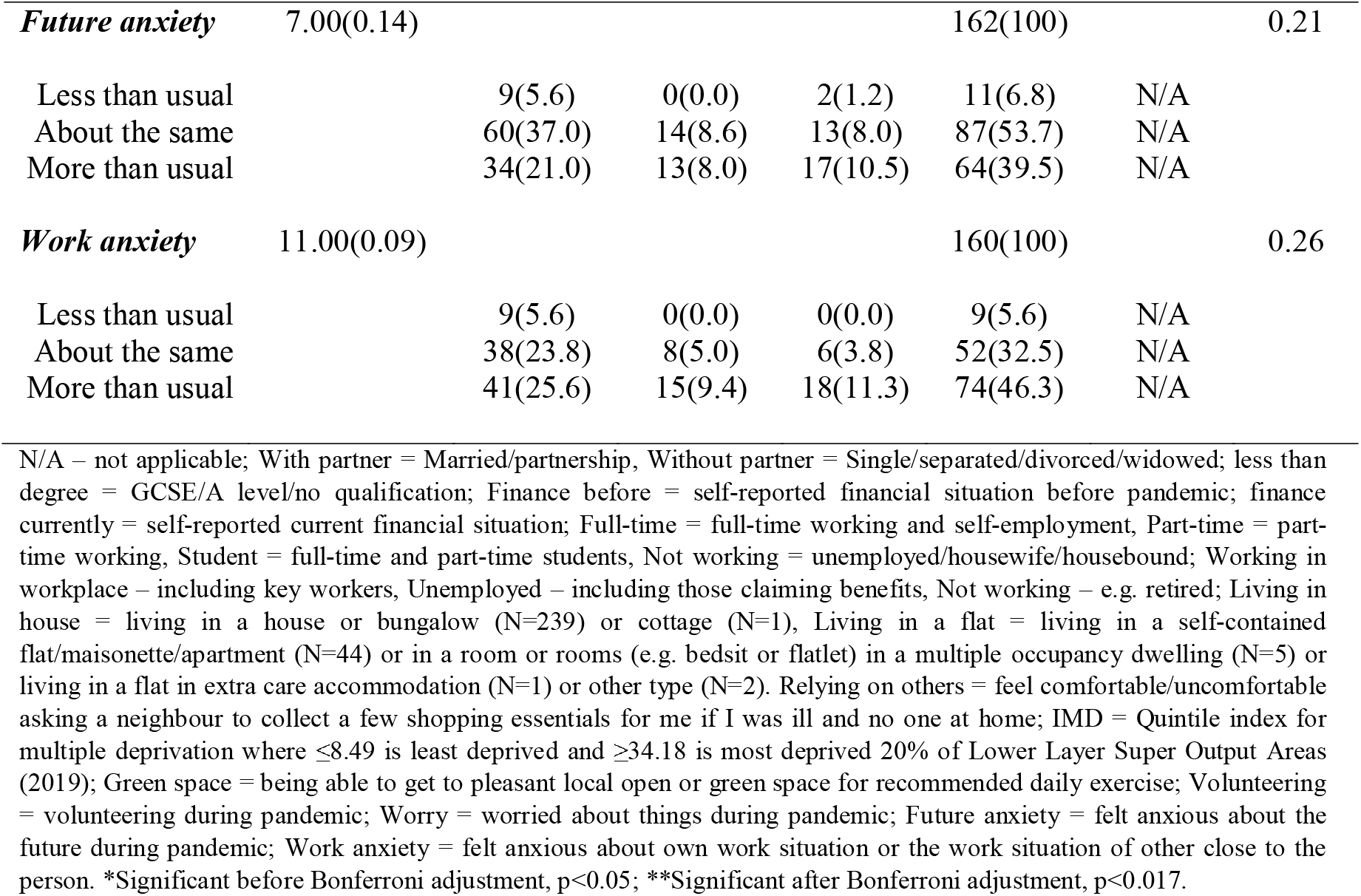
Differences between the ‘stable’, ‘volatile’, and very volatile groups. Results from the Chi-square test for independence and Fisher’s exact tests. Significant differences are bolded.

## Discussion

Adopting a resilient systems approach, the present study aimed to i) determine the proportion of people with mental stability and mental volatility during the COVID-19 pandemic in a sample of people in a city region in the North West of England and ii) establish differences between groups (stable, volatile, very volatile) in terms of individual (demographic, psychological), community, societal, and COVID-19 specific factors. Participants with low level or no change in their mental health symptoms over the course of 12 weeks were included in the ‘stable’ group reflecting their ability to adapt and cope well with the challenges of the pandemic. Associations were found between mental stability/volatility and common mental health and resilience providing validity for the use of this concept. The findings revealed that nearly two thirds of the participants (63.6%, ‘stable’ group) were coping well during the first phase of the health crisis and individual, community, societal, and COVID-19 specific mechanisms were associated with mental stability and volatility.

### Mental stability versus volatility

About one third of the participants were experiencing anxiety during the first lockdown. This is similar to reports of 32% by the ONS (2020), slightly higher than other previous UK findings of 27% ^34^, and considerably higher than pre-pandemic figures in the general UK population (5.9%) ^35^. However, this heightened level of anxiety is a normal response to an unprecedented public health crisis and lockdown measures. Whether it persists and becomes a long-term mental health problem depends upon a person’s response to the stress and their ability to cope ^38^. As the results of this study show, approximately 64% of those who completed the survey at both time points can be characterised as mentally stable, thus coping well with the pandemic and its restrictions, which is in line with previous research findings ^31,10^. This means that anxiety and depression levels remained stable over the 12 weeks for these individuals demonstrating that their mental health did not suffer during this time.

The ecological model of resilience usefully accounted for the multiple sources, within the individual, as well as their life and environment, that can impact on resilience, explaining the differences between mentally ‘stable’, ‘volatile’, and ‘very volatile’ people. Although the groups did not differ in terms of demographic and socio-demographic factors, they were different at the psychological, community, and societal levels.

#### Individual factors

At the individual level, mental stability was associated with higher level of wellbeing and mental and physical health. This link between resilience, coping, mental and physical health, and wellbeing has been clearly indicated by an extensive prior literature (e.g. ^23-25,27^). Moreover, mental stability during the pandemic was characterised by a better tolerance of uncertainty, which has also been demonstrated in an Italian sample ^31^.

#### Community factors

People have been shown to cope with the health crisis through connections with ^28^ and support from loved ones ^29^. Social connections can also affect how people respond to stress ^26^. In our study, mentally stable individuals reported feeling less socially isolated. It is known that communities are important in supporting individual resilience ^51^ and encouraging healthy coping behaviours ^52^. Sense of community, such as feeling a sense of belonging and a perception that one’s needs will be met by the community, has been associated with better mental health ^50^. Mentally stable people of this study reported their needs being met as members of their community and feeling a sense of belonging to this community.

#### Societal factors

At the societal level, those who were coping better reported having an open green space nearby which they could use to take the recommended daily exercise during lockdown. There is increasing evidence of the association between wellbeing and outdoor activity and this has also been shown to be the case during the pandemic ^28,29^.

#### COVID-specific factors

COVID-19-related anxiety has been found to hinder resilience ^31^. In the present sample, those identified as mentally volatile were characterised by excessive worry in the midst of the pandemic compared to those mentally stable who described their level of worry as ‘about the same’ as prior to the pandemic.

### Implications

These findings demonstrate that mental stability or resilience operates in an interactive way at multiple levels ^13,20^. While individual factors are important in adaptation to adversity and coping with stress, the activation of local assets to enable communities to cope with natural disasters and isolation are also necessary ^51^. Therefore, the wider determinants of health, such as the community and society we live and work in, are key to enabling the public to bounce back after adversity ^20^. Research has demonstrated that some groups in society have been disproportionately affected by the pandemic, such as ethnic minority communities ^53^ and older people ^54^ underpinned by health inequalities and unfairness. Some of the greatest drivers of health inequalities include environmental and community factors, such as extent of social isolation, unequal availability of good quality green space nearby, and differing levels of sense of community. These are also the drivers of mental health difficulties and, importantly, they are factors over which the individual can have very little control. Therefore, public health interventions should focus on developing enabling, agentic and supporting environments to facilitate people’s ability to gain control over the determinants of their health and thus promote health and well-being ^51^. The factors that were found to affect mental stability may also have an impact on people’s willingness to uptake COVID-19 vaccination. Future research, therefore, could utilise these findings to draw comparison, develop theory, and plan interventions.

### Strengths and Limitations

In addition to inviting participants from the CLAHRC NWC HHS ^43^, recruitment was extended to utilise local and social media which boosted participation in the study. This recruitment strategy ensured the recruitment of a sample from varied neighbourhoods (Table 1) including the most deprived areas of the city region, ^55^. The high levels of IMD in this sample may reflect the high levels of IMD in this region compared to the national profile, which may explain the lack of significant findings with regard to sociodemographic characteristics ^55^. Despite the extended recruitment, the week 1 survey sample was relatively small (N=290) and it was further reduced by high attrition in the 12-week follow-up (N=163). Additionally, a high proportion of the participants were women, individuals of white ethnic background (representative of the city region demographic), higher educated, and only the digitally enabled would have the capacity to complete the survey. These are common features of online surveys, which reduce the representativeness of the sample ^56^. The survey included only self-report measures, which are prone to bias. There was a very low rate of COVID-19 infections among the participants. A sample with a higher infection rate might have struggled more to cope and presented with worse mental health. Additionally, those with more mental health or other adversity, and those who have returned to work when restrictions were eased might have been less likely to stay in the study. Finally, the week 1 survey was administered 11 weeks after the nationwide lockdown began and was only able to capture two snapshots of participants’ mental health. Capturing volatility using only these two snapshots is a limitation of this study as the pattern of change in mental health in between time 1 and time 2 is unknown. Taking weekly measures would better capture mental health change.

## Conclusion

In line with previous research findings, the majority of the participants (64%) with follow-up data at week 12, were mentally stable and coping well with the challenges presented by the pandemic. This is important to record within the context of a relatively disadvantaged urban area. However, it means that the mental health of over a third of the sample during this time of unprecedented adversity was unstable. Understanding the place-based determinants that impact the volatility of mental health is key to facilitate healthy coping and the prevention of mental health problems in the population during a crisis. Attending to the wider determinants of health and developing policies that will help to create places that people can use safely and feel a part of will likely benefit the most vulnerable particularly and in doing so will reduce wellbeing inequity and improve population health.

## Data Availability

The data file used for the analyses is available from the Open Science Framework database: https://osf.io/6pwdj/

## Supporting Information

**Supplemental Table 1. All measures used in the survey. Measures from which data was used in the current study are bolded**.

**Supplemental Table 2. Recoded variables (N=290)**.

## References

1. Burn W, Mudholkar S. Impact of COVID-19 on mental health: Update from the United Kingdom. Indian J Psychiatry. 2020 Sep;62(Suppl 3):S365–S372. doi: 10.4103/psychiatry.IndianJPsychiatry_937_20.

2. Duan L, Zhu G. Psychological interventions for people affected by the COVID-19 epidemic. Lancet Psychiatry. 2020 Apr;7(4):300–302. doi: 10.1016/S2215-0366(20)30073-0.

3. Chen Q, Liang M, Li Y, Guo J, Fei D, Wang L, He L, Sheng C, Cai Y, Li X, Wang J, Zhang Z. Mental health care for medical staff in China during the COVID-19 outbreak. Lancet Psychiatry. 2020 Apr;7(4):e15–e16. doi: 10.1016/S2215-0366(20)30078-X.

4. Liem A, Wang C, Wariyanti Y, Latkin CA, Hall BJ. The neglected health of international migrant workers in the COVID-19 epidemic. Lancet Psychiatry. 2020 Apr;7(4):e20. doi: 10.1016/S2215-0366(20)30076-6.

5. Yang Y, Li W, Zhang Q, Zhang L, Cheung T, Xiang YT. Mental health services for older adults in China during the COVID-19 outbreak. Lancet Psychiatry. 2020 Apr;7(4):e19. doi: 10.1016/S2215-0366(20)30079-1. Epub 2020 Feb 19. PMID: 32085843; PMCID: PMC7128970.

6. Wang C, Pan R, Wan X, Tan Y, Xu L, Ho CS, Ho RC. Immediate Psychological Responses and Associated Factors during the Initial Stage of the 2019 Coronavirus Disease (COVID-19) Epidemic among the General Population in China. Int J Environ Res Public Health. 2020 Mar 6;17(5):1729. doi: 10.3390/ijerph17051729.

7. González-Sanguino C, Ausín B, Castellanos MÁ, Saiz J, López-Gómez A, Ugidos C, Muñoz M. Mental health consequences during the initial stage of the 2020 Coronavirus pandemic (COVID-19) in Spain. Brain Behav Immun. 2020 Jul;87:172–176. doi: 10.1016/j.bbi.2020.05.040.

8. Office for National Statistics [ONS]. Coronavirus and the social impacts on Great Britain data [Internet]. London: Office for National Statistics: 2020 [cited 20 July 2021]. Available from https://www.ons.gov.uk/peoplepopulationandcommunity/healthandsocialcare/healthandwellbeing/datasets/coronavirusandthesocialimpactsongreatbritaindata

9. Shevlin M, Butter S, McBride O, Murphy J, Gibson-Miller J, Hartman TK, Levita L, Mason L, Martinez AP, McKay R, Stocks TVA, Bennett K, Hyland P, Bentall RP. Refuting the myth of a ‘tsunami’ of mental ill-health in populations affected by COVID-19: evidence that response to the pandemic is heterogeneous, not homogeneous. Psychol Med. 2021 Apr 20:1–9. doi: 10.1017/S0033291721001665.

10. Fluharty M, Fancourt D. How have people been coping during the COVID-19 pandemic? Patterns and predictors of coping strategies amongst 26,016 UK adults. BMC Psychol. 2021 Jul 15;9(1):107. doi: 10.1186/s40359-021-00603-9.

11. Robinson, E., Sutin, A. R., Daly, M., & Jones, A. (2022). A systematic review and meta-analysis of longitudinal cohort studies comparing mental health before versus during the COVID-19 pandemic in 2020. Journal of affective disorders, 296, 567– 576. doi: 10.1016/j.jad.2021.09.098

12. Rossi NE, Bisconti TL, Bergeman CS. The role of dispositional resilience in regaining life satisfaction after the loss of a spouse. Death Stud. 2007 Nov;31(10):863–83. doi: 10.1080/07481180701603246. PMID: 17924502.

13. Windle G. What is resilience? A review and concept analysis. Reviews in Clinical Gerontology. 2011; 21(2): 152–169. doi: 10.1017/S0959259810000420

14. Bennett KM. How to achieve resilience as an older widower: Turning points or gradual change? Ageing and Society. 2010; 30(3): 369–382. doi: 10.1017/S0144686X09990572

15. Galatzer-Levy IR, Bonanno GA. Beyond normality in the study of bereavement: heterogeneity in depression outcomes following loss in older adults. Soc Sci Med. 2012 Jun;74(12):1987–94. doi: 10.1016/j.socscimed.2012.02.022.

16. Joyce S, Shand F, Tighe J, Laurent SJ, Bryant RA, Harvey SB. Road to resilience: a systematic review and meta-analysis of resilience training programmes and interventions. BMJ Open. 2018 Jun 14;8(6):e017858. doi: 10.1136/bmjopen-2017-017858.

17. Friedli L. Mental health, resilience and inequalities [Internet]. Copenhagen: World Health Organisation; 2009 [cited 30 July 2021]. Available from https://apps.who.int/iris/bitstream/handle/10665/107925/E92227.pdf

18. Windle G, Bennett KM. Resilience and caring relationships. In: Ungar M editor. The social ecologies of resilience. New York, NY: Springer; 2011. p. 219–232.

19. Ungar M. The social ecology of resilience. New York (NY): Springer; 2011.

20. Wiles JL, Wild K, Kerse N, Allen RES. Resilience from the point of view of older people: ‘There’s still life beyond a funny knee’. Soc Sci Med. 2012 Feb;74(3):416–424. doi: 10.1016/j.socscimed.2011.11.005.

21. Shapero BG, Farabaugh A, Terechina O, DeCross S, Cheung JC, Fava M, Holt DJ. Understanding the effects of emotional reactivity on depression and suicidal thoughts and behaviors: Moderating effects of childhood adversity and resilience. J Affect Disord. 2019 Feb 15;245:419–427. doi: 10.1016/j.jad.2018.11.033.

22. Loprinzi CE, Prasad K, Schroeder DR, Sood A. Stress Management and Resilience Training (SMART) program to decrease stress and enhance resilience among breast cancer survivors: a pilot randomized clinical trial. Clin Breast Cancer. 2011 Dec;11(6):364–8. doi: 10.1016/j.clbc.2011.06.008.

23. Jonker AA, Comijs HC, Knipscheer KC, Deeg DJ. The role of coping resources on change in well-being during persistent health decline. J Aging Health. 2009 Dec;21(8):1063–82. doi: 10.1177/0898264309344682.

24. Meléndez JC, Satorres E, Redondo R, Escudero J, Pitarque A. Wellbeing, resilience, and coping: Are there differences between healthy older adults, adults with mild cognitive impairment, and adults with Alzheimer-type dementia? Arch Gerontol Geriatr. 2018 Jul-Aug;77:38–43. doi: 10.1016/j.archger.2018.04.004.

25. Taylor AW, Kelly G, Dal Grande E, Kelly D, Marin T, Hey N, Burke KJ, Licinio J. Population levels of wellbeing and the association with social capital. BMC Psychol. 2017 Jul 3;5(1):23. doi: 10.1186/s40359-017-0193-0.

26. Lupe SE, Keefer L, Szigethy E. Gaining resilience and reducing stress in the age of COVID-19. Curr Opin Gastroenterol. 2020 Jul;36(4):295–303. doi: 10.1097/MOG.0000000000000646.

27. Ran L, Wang W, Ai M, Kong Y, Chen J, Kuang L. Psychological resilience, depression, anxiety, and somatization symptoms in response to COVID-19: A study of the general population in China at the peak of its epidemic. Soc Sci Med. 2020 Oct;262:113261. doi: 10.1016/j.socscimed.2020.113261.

28. Blanc J, Briggs AQ, Seixas AA, Reid M, Jean-Louis G, Pandi-Perumal SR. Addressing psychological resilience during the coronavirus disease 2019 pandemic: a rapid review. Curr Opin Psychiatry. 2021 Jan;34(1):29–35. doi: 10.1097/YCO.0000000000000665. PMID: 33230041; PMCID: PMC7751836.

29. Killgore WDS, Taylor EC, Cloonan SA, Dailey NS. Psychological resilience during the COVID-19 lockdown. Psychiatry Res. 2020 Sep;291:113216. doi: 10.1016/j.psychres.2020.113216.

30. Petzold, MB, Bendau, A, Plag, J, et al. Risk, resilience, psychological distress, and anxiety at the beginning of the COVID-19 pandemic in Germany. Brain Behav. 2020; 10:e01745. doi: 10.1002/brb3.1745

31. Panzeri A, Bertamini M, Butter S, Levita L, Gibson-Miller J, Vidotto G, Bentall RP, Bennett KM. Factors impacting resilience as a result of exposure to COVID-19: The ecological resilience model. PLoS One. 2021 Aug 18;16(8):e0256041. doi: 10.1371/journal.pone.0256041.

32. Johns Hopkins University & Medicine (Producer). Coronavirus Resource Center: 2020 [cited 03 July 2021]. Available from https://coronavirus.jhu.edu/map.html.

33. Rettie H, Daniels J. Coping and tolerance of uncertainty: Predictors and mediators of mental health during the COVID-19 pandemic. Am Psychol. 2021 Apr;76(3):427–437. doi: 10.1037/amp0000710.

34. Dawson DL, Golijani-Moghaddam N. COVID-19: Psychological flexibility, coping, mental health, and wellbeing in the UK during the pandemic. J Contextual Behav Sci. 2020 Jul;17:126–134. doi: 10.1016/j.jcbs.2020.07.010.

35. McManus S, Bebbington PE, Jenkins R, Brugha T. Mental health and wellbeing in England: Adult Psychiatric Morbidity Survey 2014. Leeds: NHS Digital, 2016.

36. Cheng SK, Wong CW, Tsang J, Wong KC. Psychological distress and negative appraisals in survivors of severe acute respiratory syndrome (SARS). Psychol Med. 2004 Oct;34(7):1187–95. doi: 10.1017/s0033291704002272.

37. Sprang G, Silman M. Posttraumatic stress disorder in parents and youth after health-related disasters. Disaster Med Public Health Prep. 2013 Feb;7(1):105–10. doi: 10.1017/dmp.2013.22.

38. PeConga EK, Gauthier GM, Holloway A, Walker RSW, Rosencrans PL, Zoellner LA, Bedard-Gilligan M. Resilience is spreading: Mental health within the COVID-19 pandemic. Psychol Trauma. 2020 Aug;12(S1):S47-S48. doi: 10.1037/tra0000874.

39. Galatzer-Levy IR, Huang SH, Bonanno GA. Trajectories of resilience and dysfunction following potential trauma: A review and statistical evaluation. Clin Psychol Rev. 2018 Jul;63:41–55. doi: 10.1016/j.cpr.2018.05.008.

40. Fluharty M, Bu F, Steptoe A, Fancourt D. Coping strategies and mental health trajectories during the first 21 weeks of COVID-19 lockdown in the United Kingdom. Soc Sci Med. 2021 Jun;279:113958. doi: 10.1016/j.socscimed.2021.113958.

41. Ogueji IA, Okoloba MM, Demoko Ceccaldi BM. Coping strategies of individuals in the United Kingdom during the COVID-19 pandemic. Curr Psychol. 2021 Jan 3:1–7. doi: 10.1007/s12144-020-01318-7.

42. Shevlin M, McBride O, Murphy J, Miller JG, Hartman TK, Levita L, Mason L, Martinez AP, McKay R, Stocks TVA, Bennett KM, Hyland P, Karatzias T, Bentall RP. Anxiety, depression, traumatic stress and COVID-19-related anxiety in the UK general population during the COVID-19 pandemic. BJPsych Open. 2020 Oct 19;6(6):e125. doi: 10.1192/bjo.2020.109.

43. Giebel C, McIntyre JC, Alfirevic A, Corcoran R, Daras K, Downing J, Gabbay M, Pirmohamed M, Popay J, Wheeler P, Holt K, Wilson T, Bentall R, Barr B. The longitudinal NIHR ARC North West Coast Household Health Survey: exploring health inequalities in disadvantaged communities. BMC Public Health. 2020 Aug 18;20(1):1257. doi: 10.1186/s12889-020-09346-5.

44. Kroenke K, Spitzer RL, Williams JB. The PHQ-9: validity of a brief depression severity measure. J Gen Intern Med. 2001 Sep;16(9):606–13. doi: 10.1046/j.1525-1497.2001.016009606.x

45. Spitzer RL, Kroenke K, Williams JB, Löwe B. A brief measure for assessing generalized anxiety disorder: the GAD-7. Arch Intern Med. 2006 May 22;166(10):1092–7. doi: 10.1001/archinte.166.10.1092.

46. Tennant R, Hiller L, Fishwick R, Platt S, Joseph S, Weich S, Parkinson J, Secker J, Stewart-Brown S. The Warwick-Edinburgh Mental Well-being Scale (WEMWBS): development and UK validation. Health Qual Life Outcomes. 2007 Nov 27;5:63. doi: 10.1186/1477-7525-5-63.

47. Smith BW, Dalen J, Wiggins K, Tooley E, Christopher P, Bernard J. The brief resilience scale: assessing the ability to bounce back. Int J Behav Med. 2008;15(3):194–200. doi: 10.1080/10705500802222972.

48. Carleton RN, Norton MA, Asmundson GJ. Fearing the unknown: a short version of the Intolerance of Uncertainty Scale. J Anxiety Disord. 2007;21(1):105–17. doi: 10.1016/j.janxdis.2006.03.014.

49. Freeston M, Rhéaume J, Letarte H, Dugas MJ, Ladouceur R. Why do people worry? Personality & Individual Differences. 1994; 17:791–802. doi: 10.1016/0191-8869(94)90048-5

50. Peterson NA, Speer PW, McMillan DW. Validation of A brief sense of community scale: Confirmation of the principal theory of sense of community. Journal of Community Psychology. 2008; 36(1):61–73. doi: 10.1002/jcop.20217

51. WHO, World Health Organization Regional Office for Europe. Strengthening resilience: a priority shared by Health 2020 and the Sustainable Development goals [Internet]. Copenhagen: World Health Organization Regional Office for Europe; 2017 [cited 18 June 2019]. Available from: http://www.euro.who.int/data/assets/pdf_file/0005/351284/resilience-report-20171004-h1635.pdf

52. Sippel LM, Pietrzak, RH, Charney DS, Mayes LC, Southwick SM. How does social support enhance resilience in the trauma-exposed individual? Ecology and Society. 2015; 20(4):10. doi: 10.5751/ES-07832-200410

53. PHE, Public Health England. Beyond the data: Understanding the impact of COVID-19 on BAME groups [Internet]. London: Public Health England; 2020 [cited 12 Jan 2020]. Available from https://assets.publishing.service.gov.uk/government/uploads/system/uploads/attachment_data/file/892376/COVID_stakeholder_engagement_synthesis_beyond_the_data.pdf

54. Martins Van Jaarsveld G. The Effects of COVID-19 Among the Elderly Population: A Case for Closing the Digital Divide. Front Psychiatry. 2020 Nov 12;11:577427. doi: 10.3389/fpsyt.2020.577427.

55. Public Health Institute (PHI). Vulnerable individuals and groups profile Liverpool City Region [Internet]. Liverpool: Public Health Institute, Faculty of Health, John Moores University; 2021 [cited 12 Jan 2022]. Available from: https://www.ljmu.ac.uk/~/media/phi-reports/pdf/2021-03-vulnerable-groups-profile-liverpool-city-region.pdf

56. Ball HL. Conducting Online Surveys. Journal of Human Lactation [Internet]. 2019 Aug [cited 2022 Mar 4];35(3):413–7. Available from: https://search-ebscohost-com.liverpool.idm.oclc.org/login.aspx?direct=true&db=jlh&AN=137411705&site=eds-live&scope=site

